# DNA Methylation Architecture of the *ACE2* gene in Nasal Cells

**DOI:** 10.1101/2020.08.25.20182105

**Authors:** Andres Cardenas, Sheryl L. Rifas-Shiman, Joanne E. Sordillo, Dawn L. DeMeo, Andrea A Baccarelli, Marie-France Hivert, Diane R Gold, Emily Oken

## Abstract

Severe Acute Respiratory Syndrome Coronavirus 2 (SARS-CoV-2) has led to the global coronavirus disease 2019 (COVID-19) pandemic. SARS-CoV-2 enters cells via angiotensin-Converting Enzyme 2 (ACE2) receptors, highly expressed in nasal epithelium with parallel high infectivity.^1,2^ The nasal epigenome is in direct contact with the environment and could explain COVID-19 disparities by reflecting social and environmental influences on *ACE2* regulation. We collected nasal swabs from anterior nares of 547 children, measured DNA methylation (DNAm), and tested differences at 15 *ACE2* CpGs by sex, age, race/ethnicity and epigenetic age. *ACE2* CpGs were differentially methylated by sex with 12 sites having lower DNAm (mean=12.71%) and 3 sites greater DNAm (mean=1.45%) among females relative to males. We observed differential DNAm at 5 CpGs for Hispanic females (mean absolute difference=3.22%) and lower DNAm at 8 CpGs for Black males (mean absolute difference=1.33%), relative to white participants. Longer DNAm telomere length was associated with greater *ACE2* DNAm at 11 and 13 CpGs among males (mean absolute difference=7.86%) and females (mean absolute difference=8.21%), respectively. Nasal *ACE2* DNAm differences could contribute to our understanding COVID-19 severity and disparities reflecting upstream environmental and social influences.

## Introduction

The global COVID-19 pandemic caused by SARS-CoV-2 has infected over 29 million individuals and claimed over 925,000 lives worldwide as of September 15, 2020.^3^ The transmission of SARS-CoV-2 is thought to mostly occur via respiratory droplets and aerosol, both targeting the respiratory system.^4,5^ The SARS-CoV-2 spike (S) glycoprotein allows viral entry into the cell by binding to the extracellular domain of the ACE2 receptor.^6,7^ It is thought that ACE2 is necessary for infection by SARS-CoV-2 and that it contributes to disease severity.^8^ In a different strain of SARS coronavirus, transgenic mice expressing human ACE2 in airway and other epithelia cells develop a rapidly lethal infection.^9^

The nasal epithelium, in direct contact with the environment, is a primary target of SARS-CoV-2 infection. Both ACE2 expression and SARS-CoV-2 infectivity are higher in nasal epithelium relative to the lungs.^1^ The nose is exposed to environmental challenges such as air pollution, cigarette smoke, and allergens that could modulate SARS-CoV-2 infectivity and COVID-19 severity and may explain disparities. For example, it has been hypothesized that exposure to air pollution and cigarette smoke could worsen immune response to SARS-CoV-2.^10,11^ This is in part supported by studies demonstrating that ACE2 expression is increased after PM_2.5_ exposure^12,13^ and among current smokers.^8,14^ Transcriptional regulation via DNAm of ACE2 has been documented in peripheral leukocytes and T cells^15,16^ and hypothesized to modulate SARS-CoV-2 infectivity and COVID-19 severity.^17,18^

The *ACE* gene is located in the X-chromosome and shown to escape X-inactivation in females^18^ with heterogeneous but significant male-bias in expression in several tissues.^19^ However, no study has examined the DNAm landscape of the *ACE2* gene in target nasal cells. Identifying demographic and biological differences could help elucidate disparities and the heterogeneity observed in COVID-19 cases.

## RESULTS

A total of 277 males (50.6%) and 270 females (49.4%) provided nasal samples at a mean age of 12.95 years (standard deviation [SD] 0.65; range 11.8 to 15.4 years). A total of 367 participants were white (67.1%), 89 Black (16.3%), 23 Hispanic (4.2%), 17 Asian (3.1%) and 51 (9.3%) more than one race. Mean (SD) for estimated DNAm telomere length and epigenetic age of the skin and blood clock were 7.11 Kb (0.38) and 15.7 years (1.94), **Table S1**.

We used the Illumina EPIC array that estimates DNAm at 16 CpG sites annotated to the *ACE2* gene. These sites cover regions between 1,500-200 and 200-0 bases of the start site, exon, body and 3’ UTR regions of the *ACE2* gene. Six CpGs were annotated to a Transcription Factor Binding Site (TFBS) with relatively high evidence from 4-6 experiments extracted from Encyclopedia of DNA Elements (ENCODE) supporting transcription binding at that region. After excluding one CpG due to low detection, DNAm levels of the other 15 CpGs ranged from 2.53% to 95.47% and 14 sites had DNAm levels above 60% with robust significant differences by sex (FDR<0.003), **Table 1 and Figure 1-A**. After adjusting for age, 12 of the 15 CpGs in the *ACE2* gene had significantly lower DNAm levels in females relative to males ranging from −31.56% at cg04013915 to −0.47% at cg18877734, **Table 2**. Three CpGs (cg18458833, cg08559914 and cg05039749) showed greater DNAm in females relative to males; these were also the only sites negatively correlated with DNAm of other CpGs. Most CpGs were strongly positively correlated (*r_S_*>0.5) except for cg05039749, annotated to the body of the gene towards the 3’ UTR, showing moderate negative correlations with all other sites, **Figure 1-B**. Given large differences by sex and *ACE2* escaping X-inactivation, we analyzed age associations stratified by sex and observed that chronological age was associated with consistently lower but weak DNAm among females and only 4 CpGs reached unadjusted significance (*p*<0.05); associations did not survive false discovery adjustment. No associations with age were observed among males, **Table S2**. However, power was limited by the narrow age range.

**Table 1.** Genomic annotation of probes located within the Illumina EPIC array and nasal DNA methylation descriptive statistics for the angiotensin-converting enzyme (*ACE2*) gene located in the X-chromosome and sex differences.

| CpG Name | position | Gene Group | TFBS coordinates<br>X-chromosomes | TFBS<br>evidence count | Mean (SD) %-DNA methylation |  |  | †Adjusted %-<br>DNAm difference |
| --- | --- | --- | --- | --- | --- | --- | --- | --- |
|  |  |  |  |  | Overall | Females | Males |  |
| cg18458833 | 15621477 | TSS1500 |  |  | 95.47 (1.84) | 96.28 (0.70) | 94.68 (1.84) | 1.52 |
| cg21598868 | 15621167 | TSS1500 |  |  | 87.88 (2.51) | 81.56 (6.08) | 94.05 (2.51) | -12.12 |
| cg18877734 | 15621084 | TSS1500 |  |  | 94.26 (2.22) | 94.00 (2.14) | 94.51 (2.22) | -0.45 |
| cg03536816 | 15621042 | TSS1500;Body |  |  | 66.55 (12.88) | 56.14 (12.14) | 76.69 (12.88) | -21.29 |
| cg08559914 | 15620240 | TSS200 |  |  | 91.80 (4.84) | 92.98 (2.88) | 90.65 (4.84) | 1.84 |
| cg16734967 | 15620103 | 1stExon;5'UTR | 15619908-15620856 | 6 | 77.51 (8.28) | 72.46 (9.57) | 82.44 (8.28) | -9.76 |
| cg10408040 | 15619136 | 5'UTR;ExonBnd |  |  | 92.76 (1.30) | 91.05 (2.75) | 94.42 (1.30) | -3.05 |
| cg05241917 | 15616197 | Body | 15616057-15616589 | 4 | 91.16 (4.48) | 89.78 (4.90) | 92.49 (4.48) | -2.31 |
| cg20119767 | 15612969 | ExonBnd;Body | 15612415-15612981 | 6 | 83.72 (5.01) | 79.52 (4.90) | 87.82 (5.01) | -8.67 |
| cg04013915 | 15586143 | Body | 15586103-15586918 | 6 | 67.24 (10.23) | 51.64 (10.36) | 82.44 (10.23) | -31.47 |
| cg16716680 | 15585368 | Body |  |  | 76.87 (5.76) | 66.82 (6.54) | 86.68 (5.76) | -20.08 |
| cg25176872 | 15584988 | Body | 15582921-15585025 | 6 | 89.64 (2.41) | 85.20 (4.37) | 93.97 (2.41) | -8.49 |
| cg22422976 | 15584867 | Body | 15582921-15585025 | 6 | 81.54 (2.42) | 68.72 (4.97) | 94.04 (4.42) | -25.22 |
| cg05039749 | 15583512 | Body |  |  | 2.53 (0.45) | 3.16 (1.47) | 1.91 (0.45) | 1.00 |
| cg23232263 | 15579482 | 3'UTR |  |  | 90.23 (3.72) | 85.52 (5.73) | 94.81 (3.72) | -8.96 |

†Differences in DNAm for females relative to males adjusted by age from a multivariate regression models all results were significant
at FDR<0.003

**Figure 1-A.**
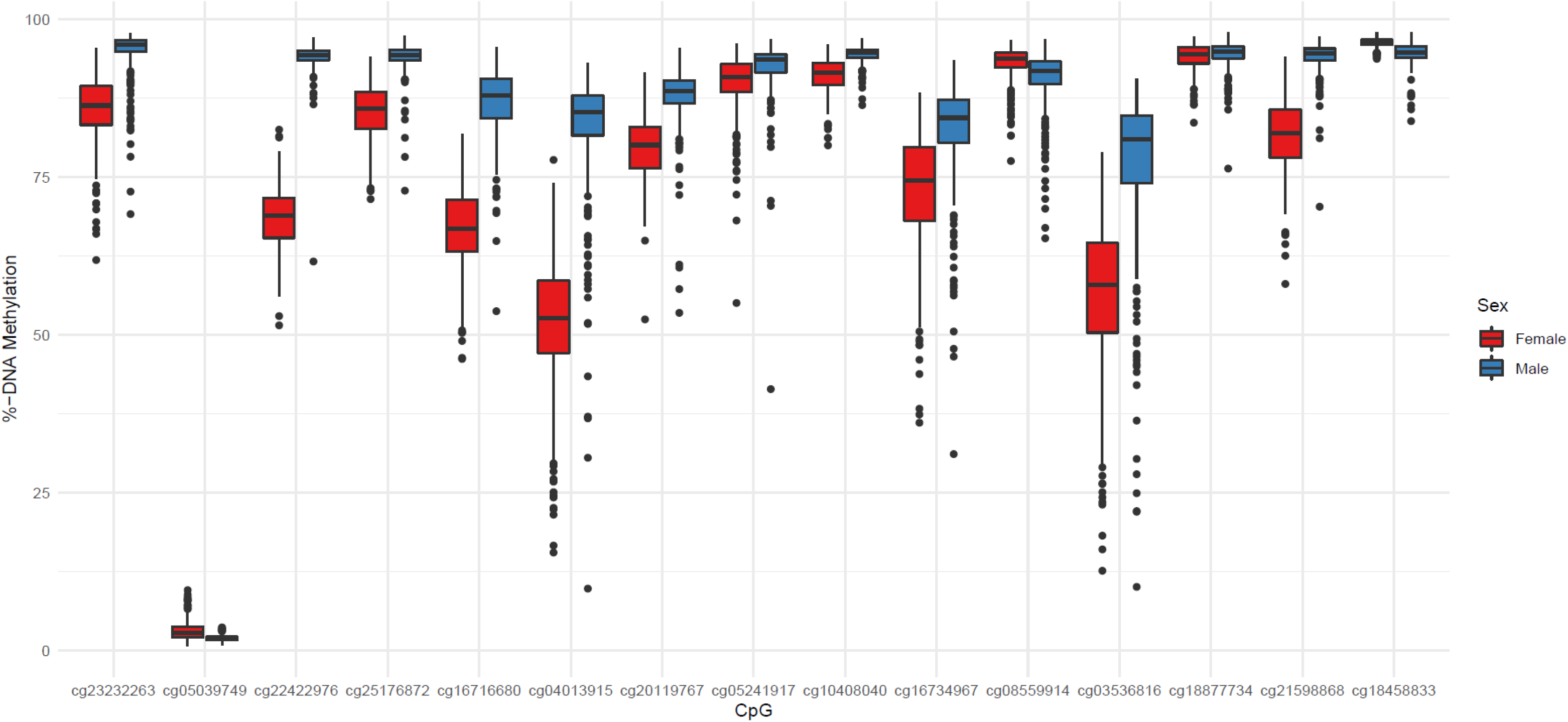
Boxplots of DNA methylation in nasal cells for the angiotensin-converting enzyme 2 (*ACE2*) gene by sex among 15 CpG sites

**Table 2.** Differences in %-DNAm in Black and Hispanic relative to white participants adjusted for age and stratified by sex.

|  | Females |  |  |  |  |  |  | Males |  |  |  |  |  |  |  |
| --- | --- | --- | --- | --- | --- | --- | --- | --- | --- | --- | --- | --- | --- | --- | --- |
| CpG | Black |  |  |  | Hispanic |  |  |  | Black |  |  |  | Hispanic |  |  |
|  | %-DNAm difference | *p* | FDR |  | %-DNAm difference | *p* | FDR |  | %-DNAm difference | *p* | FDR |  | %-DNAm difference | *p* | FDR |
| cg18458833 | -0.16 | 0.18 | 0.33 |  | 0.11 | 0.49 | 0.57 |  | **-0.75** | **0.0011** | **0.005** |  | 0.31 | 0.49 | 0.90 |
| cg21598868 | 1.76 | 0.03 | 0.23 |  | 2.45 | 0.11 | 0.20 |  | -0.47 | 0.07 | 0.10 |  | 0.34 | 0.41 | 0.90 |
| cg18877734 | 0.47 | 0.09 | 0.30 |  | 0.60 | 0.32 | 0.40 |  | **-0.77** | **0.0036** | **0.011** |  | 0.20 | 0.73 | 0.96 |
| cg03536816 | 0.28 | 0.88 | 0.94 |  | **6.63** | **0.0051** | **0.019** |  | -1.92 | 0.20 | 0.22 |  | -0.28 | 0.92 | 0.96 |
| cg08559914 | -0.71 | 0.10 | 0.30 |  | **1.12** | **2.42x10<sup>-5</sup>** | **0.00036** |  | **-1.76** | **0.0015** | **0.006** |  | 0.98 | 0.34 | 0.90 |
| cg16734967 | -1.54 | 0.30 | 0.51 |  | **4.44** | **0.0039** | **0.019** |  | **-3.44** | **4.46x10<sup>-4</sup>** | **0.005** |  | 1.26 | 0.35 | 0.90 |
| cg10408040 | 0.45 | 0.18 | 0.33 |  | **-1.69** | **0.0012** | **0.009** |  | -0.36 | 0.04 | 0.07 |  | 0.09 | 0.65 | 0.96 |
| cg05241917 | 0.16 | 0.77 | 0.94 |  | 0.99 | 0.21 | 0.31 |  | **-1.36** | **7.18x10<sup>-4</sup>** | **0.005** |  | 0.85 | 0.14 | 0.90 |
| cg20119767 | 0.04 | 0.96 | 0.96 |  | 1.92 | 0.27 | 0.37 |  | **-1.30** | **0.02** | **0.045** |  | 0.11 | 0.91 | 0.96 |
| cg04013915 | 1.01 | 0.52 | 0.78 |  | 3.32 | 0.05 | 0.10 |  | -1.66 | 0.10 | 0.13 |  | 0.70 | 0.54 | 0.90 |
| cg16716680 | 2.07 | 0.029 | 0.23 |  | 1.17 | 0.55 | 0.58 |  | -0.86 | 0.27 | 0.27 |  | -1.69 | 0.24 | 0.90 |
| cg25176872 | 1.26 | 0.08 | 0.30 |  | 2.66 | 0.024 | 0.06 |  | **-0.64** | **0.006** | **0.016** |  | -0.02 | 0.96 | 0.96 |
| cg22422976 | 1.12 | 0.17 | 0.33 |  | 2.6 | 0.15 | 0.25 |  | -0.28 | 0.20 | 0.22 |  | 0.02 | 0.89 | 0.96 |
| cg05039749 | 0.04 | 0.86 | 0.94 |  | 0.04 | 0.90 | 0.90 |  | 0.12 | 0.10 | 0.13 |  | -0.08 | 0.54 | 0.90 |
| cg23232263 | -0.18 | 0.83 | 0.94 |  | **2.21** | **0.014** | **0.042** |  | **-0.60** | **0.016** | **0.03** |  | 0.21 | 0.54 | 0.90 |

**Figure 1-B.**
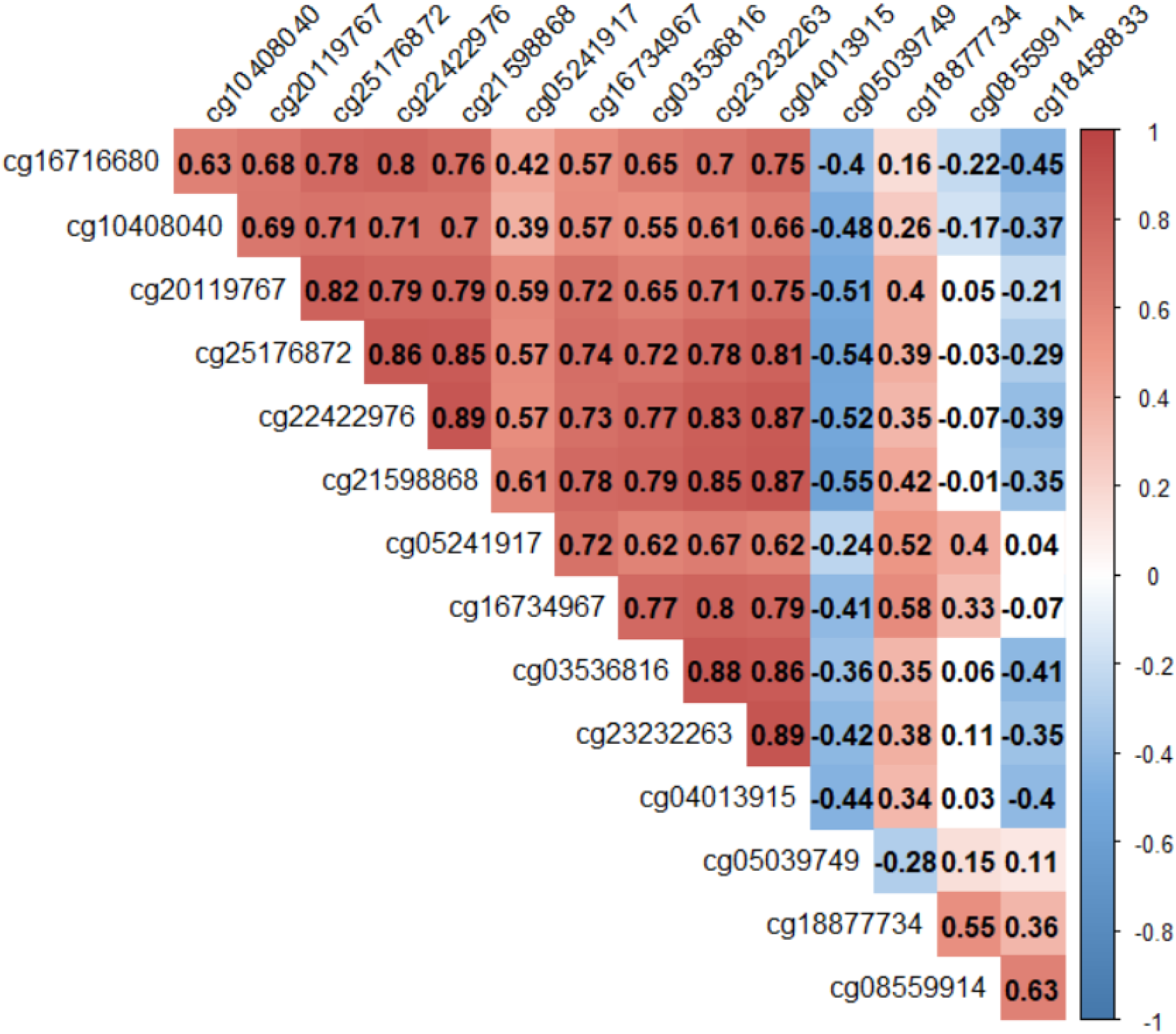
Spearman correlation coefficients of nasal cell DNA methylation levels among CpGs found in the angiotensin-converting enzyme (*ACE2*) gene.

Hispanic females had significantly greater DNAm of 6.63% at cg03536816, 1.12% at cg08559914, 4.44% at cg16734967 and 2.21% at cg23232263 while cg10408040 showed a 1.69% lower DNAm relative to white females (FDR<0.05), **Table 2**. A local *ACE2* gene Manhattan plot for the association with Hispanic females with genomic annotation and correlation structure is shown in **Figure 1-C**. Among Black males, significantly lower DNAm at 8 CpG sites was present relative to white males. Specifically, lower DNAm of 0.75% at cg18458833, 0.77% at cg18877734, 1.76% at cg08559914, 3.44% at cg16734967, 1.36% at cg05241917, 1.30% at cg20119767, 0.64% at cg25176872 and 0.60% at cg23232263 for Black relative to white males was observed, adjusting for age (FDR<0.05), **Table 2**. A local *ACE2* gene Manhattan plot for the association among Black males with genomic annotation and correlation structure is presented in **Figure 1-D**. No differences were observed among Asian or participants of more than one race, **Table S2**.

**Figure 1-C.**
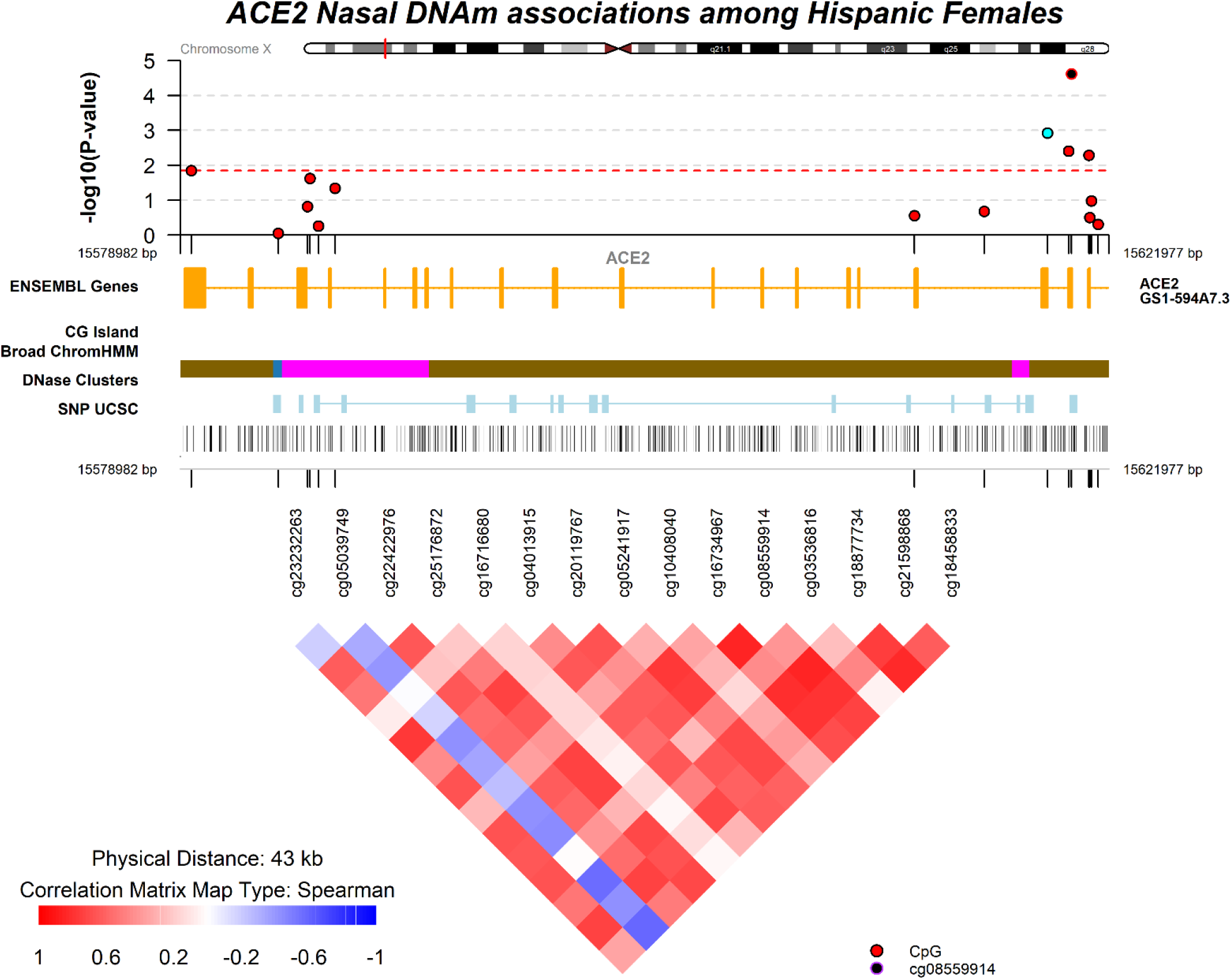
Local Manhattan plot for the association of *ACE2* DNA methylation for Hispanic relative to white females. Spearman correlation matrix for CpGs among all females.

**Figure 1-D.**
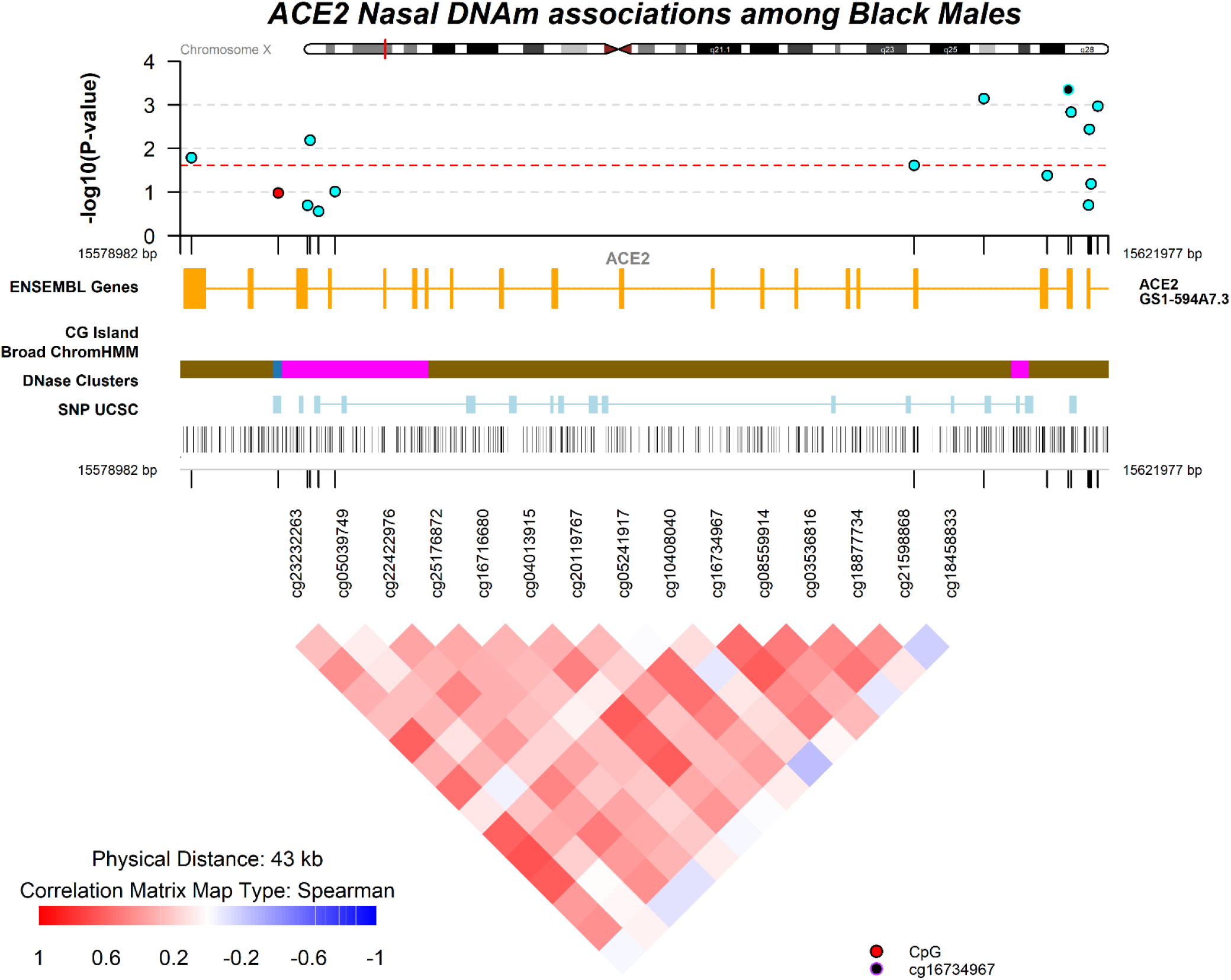
Local Manhattan plot for the association of *ACE2* DNA methylation for Black relative to white males. Spearman correlation matrix for CpGs among all males.

DNAm telomere length (DNAmTL) and the Skin and Blood Epigenetic clock were chosen as markers of epigenetic aging as they performed relatively well against chronological age, **Figure S1**. Longer DNAm telomere length adjusted for age (DNAmTLadjAge) was consistently and strongly associated with greater *ACE2* DNAm for males with a mean increase of 7.86% increased across 11 CpGs. For females a mean of 8.21% greater DNAm across 13 CpGs was observed. Weaker positive relationships were observed for 5 CpGs with epigenetic age acceleration of the skin and blood clock, **Table 3**.

**Table 3.** *Differences in %-DNAm of the *ACE2* gene in nasal cells by estimated age-adjusted DNA methylation telomere length residuals (DNAmTLAdjAge) and epigenetic age acceleration of the skin and blood clock stratified by sex

| CpG | DNAmTLAdjAge |  |  |  |  |  | Skin & Blood Clock Acceleration |  |  |  |  |  |
| --- | --- | --- | --- | --- | --- | --- | --- | --- | --- | --- | --- | --- |
|  | Females |  |  | Males |  |  | Females |  |  | Males |  |  |
|  | %-DNAm difference | *p* | FDR | %-DNAm difference | *P* | FDR | %-DNAm difference | *p* | FDR | %-DNAm difference | *p* | FDR |
| cg18458833 | 0.06 | 0.57 | 0.61 | 0.53 | 0.06 | 0.08 | -0.03 | 0.16 | 0.18 | -0.01 | 0.86 | 0.92 |
| cg21598868 | **6.74** | **1.24x10<sup>-11</sup>** | **2.32x10<sup>-11</sup>** | **3.17** | **6.13x10<sup>-19</sup>** | **1.31x10<sup>-18</sup>** | 0.39 | 0.05 | 0.08 | -0.01 | 0.92 | 0.92 |
| cg18877734 | **2.13** | **1.06x10<sup>-10</sup>** | **1.77x10<sup>-10</sup>** | **2.48** | **4.43x10<sup>-8</sup>** | **7.39x10<sup>-8</sup>** | 0.15 | 0.05 | 0.08 | 0.05 | 0.51 | 0.92 |
| cg03536816 | **24.34** | **5.06x10<sup>-124</sup>** | **7.59x10<sup>-123</sup>** | **28.2** | **6.50x10<sup>-53</sup>** | **9.74x10<sup>-52</sup>** | **0.97** | **0.02** | **0.05** | 0.15 | 0.72 | 0.92 |
| cg08559914 | **4.44** | **2.14x10<sup>-25</sup>** | **6.43x10<sup>-25</sup>** | **7.41** | **8.60x10<sup>-27</sup>** | **4.30x10<sup>-26</sup>** | 0.19 | 0.03 | 0.06 | -0.03 | 0.83 | 0.92 |
| cg16734967 | **17.32** | **2.87x10<sup>-50</sup>** | **1.08x10<sup>-49</sup>** | **11.68** | **1.59x10<sup>-19</sup>** | **3.97x10<sup>-19</sup>** | **0.92** | **0.015** | **0.05** | 0.04 | 0.86 | 0.92 |
| cg10408040 | 0.16 | 0.71 | 0.71 | 0.27 | 0.16 | 0.18 | 0.2 | 0.02 | 0.06 | -0.05 | 0.21 | 0.80 |
| cg05241917 | **6.59** | **9.62x10<sup>-15</sup>** | **2.40 x10<sup>-14</sup>** | **4.18** | **1.77x10<sup>-28</sup>** | **1.33x10<sup>-27</sup>** | **0.37** | **0.004** | **0.03** | -0.08 | 0.43 | 0.92 |
| cg20119767 | **2.12** | **0.0003** | **0.0004** | **2.48** | **0.0027** | **0.0037** | **0.44** | **0.008** | **0.04** | -0.25 | 0.07 | 0.80 |
| cg04013915 | **19.83** | **2.31x10<sup>-72</sup>** | **1.16x10<sup>-71</sup>** | **18.2** | **7.48x10<sup>-20</sup>** | **2.24x10<sup>-19</sup>** | **1.15** | **0.004** | **0.03** | 0.04 | 0.88 | 0.92 |
| cg16716680 | **4.15** | **5.52x10<sup>-6</sup>** | **7.52x10<sup>-6</sup>** | 0.58 | 0.52 | 0.52 | 0 | 0.98 | 0.98 | 0.18 | 0.20 | 0.80 |
| cg25176872 | **2.32** | **0.0013** | **0.0015** | **1.08** | **9.18x10<sup>-5</sup>** | **0.00014** | 0.22 | 0.16 | 0.18 | -0.01 | 0.86 | 0.92 |
| cg22422976 | **5.03** | **3.67x10<sup>-13</sup>** | **7.86x10<sup>-13</sup>** | **1.55** | **9.13x10<sup>-13</sup>** | **1.71x10<sup>-12</sup>** | 0.18 | 0.36 | 0.38 | -0.08 | 0.15 | 0.80 |
| cg05039749 | **0.72** | **7.00x10<sup>-7</sup>** | **1.05x10<sup>-6</sup>** | -0.08 | 0.25 | 0.27 | 0.09 | 0.03 | 0.06 | 0.01 | 0.57 | 0.92 |
| cg23232263 | **11.04** | **4.50Ex10<sup>-88</sup>** | **3.37x10<sup>-87</sup>** | **6.05** | **7.67x10<sup>-25</sup>** | **2.87x10<sup>-24</sup>** | 0.37 | 0.08 | 0.11 | 0.06 | 0.38 | 0.92 |

\*Adjusted for race/ethnicity

## DISCUSSION

In summary, all *ACE2* CpGs tested differed strongly and significantly by sex with most sites showing a decrease in DNAm among females relative to males. In sex stratified analyses, moderate greater DNAm for Hispanic females and consistently lower DNAm among Black males relative to white participants was observed. Longer telomere length adjusted for age was strongly associated with greater *ACE2* DNAm and this was consistent among males and females across most CpGs.

The *ACE2* gene is located in the X-chromosome and escapes X-inactivation in females with potential tissue specificity and heterogenous male expression bias, partially attributed to sex steroids.^20^ This heterogeneity has been documented in data from an RNAseq Atlas identifying co-expression of both *ACE2* and the accessory proteases among subsets of respiratory epithelial cells as putative targets of SARS-CoV-2 infection. Specifically, nasal goblet cells and ciliated cells comprised the highest fraction of cells expressing both ACE2 and the viral entry associated protease TMPRSS2.^2,21^ The relatively high ACE2 expression in the nasal epithelium its coupled with high infectivity in nasal epithelium with progressively lower expression and infectivity in the bronchial and alveolar regions.^1,22^ Increased ACE2 expression has been hypothesized to partially explain differences in COVID-19 severity.^23^ For example, lung transcriptome data from patients with comorbid conditions associated with increased risk of severe COVID-19 like hypertension, diabetes, and chronic obstructive lung disease show increased ACE2 expression.^24^

ACE2 expression has been reported to be higher in Asians compared with white and African Americans from type II alveolar cells.^25^ However, other reports have found no difference in ACE2 expression by race.^26^ Data from lung tissue has shown lower DNAm in females relative to males at cg23232263 and cg16734967 consistent with our finding for females using nasal cells.^27^ In addition, a strong negative association between chronological age and DNAm of cg08559914 from airway epithelial cells was reported. This was not confirmed in our data as associations with age were not observed.^27^ However, expression of ACE2 in the lung has been demonstrated to increase with age.^26^ In nasal samples, lower ACE2 expression among children relative to older adults has been reported.^28^ Age associated expression differences are hypothesized to partially explain the relatively mild COVID-19 documented cases in children. Yet, large racial inequalities in need for hospitalization among children have been reported. Data from the U.S. shows that hospitalizations for Hispanic and Black children are much more common relative to white children.^29^ Our results support the hypothesis that *ACE2* hypomethylation in nasal epithelium among black males could lead to increased SARS-CoV-2 infectivity and COVID-19 severity via greater abundance of ACE2 receptors. Results among females need to be cautiously interpreted given ACE2 escape from X-inactivation. *ACE2* genetic variants are rare, not shown to differ by sex or populations.^30,31^ Therefore, we hypothesize that observed DNA methylation variation originates from social and environmental exposures which include social and environmental racism.

Our study has some limitations. We do not have ACE2 expression which would inform the underlying hypothesis of epigenetic regulation. We hypothesize that the relationship between DNAm, expression and ACE2 receptors is likely complex and depends on the genomic context, cells and life-stage. The absence of age-related DNAm changes could be due to the narrow age range and young profile of our study. However, measures of epigenetic age acceleration might be more sensitive to age-related physiological changes among children as reflected in strong associations with DNAm telomere length. Although most participants in our cohort were white, the relatively large sample allowed us to test differences stratifying by sex and test differences by race/ethnicity.

The epigenome is at the interface of the environment and genomic regulation. Nasal cells which are directly in contact with the environment have been shown to play a key role in SARS-CoV-2 infection. Our results demonstrate that *ACE2* nasal DNAm reflects differences by sex, race/ethnicity and biological aging. The nasal epigenetic architecture of the *ACE2* gene could contribute to our understanding of COVID-19 and environmental disparities.

### Materials and Methods

#### Study Population

We included participants from Project Viva, a prospective pre-birth cohort from eastern Massachusetts recruited between 1999 and 2002 during the initial prenatal visit at Atrius Harvard Vanguard Medical Associates.^32^ Eligibility criteria for cohort recruitment included fluency in English, gestational age <22 weeks at the first prenatal visit, and singleton pregnancy. At the early childhood visit, we asked the mother to choose one or more racial/ethnic groups for her child. We created five racial/ethnic groups based on the mother’s response including white, Black, Hispanic, Asian, and other (>1 race or other). We used mother’s race to fill in missing child race values. Of the total 2128 live births, 547 children were re-contacted during an early-teen in-person visit at a mean age 12.9 years (range; 11.8 to 15.4 y) and provided consent for nasal swab sample collection and epigenetic studies. Mothers provided written informed consent at recruitment and at postpartum follow-up visits. Written informed consent was obtained from parent and/or legal guardian of minors and children provided written assent. Institutional Review Board of Harvard Pilgrim Health Care reviewed and approved all study protocols. All methods were carried out in accordance with relevant guidelines and regulations stipulated by the institutional review board.

#### Nasal DNA methylation analyses

Trained research staff collected nasal swabs from the anterior nares, previously shown to yield respiratory epithelial cells.^33^ Nasal swabs were immediately stored in lysis buffer and frozen until processing. We isolated DNA using the Maxwell 16 Buccal Swab LEV DNA Purification Kit following the manufacturer’s instructions (Promega, Madison, WI, USA). After DNA extraction from the nasal samples we measured DNA methylation with the Infinium MethylationEPIC BeadChip. All DNAm data were imported into the *R* statistical software and samples were excluded based quality control check as previously described.^34^ We preprocessed all of the DNA methylation data using functional normalization to adjust for technical variability^35^ and adjusted for probe-type variability using RCP.^36^ Lastly, we used ComBat from the *sva* package to adjust for sample plate (seven 96-well plates) as a technical variable.^37^ One CpG site of the *ACE2* gene, cg05748796, was excluded as it failed detection in 143 of the samples (*p*>0.05). The other 15 CpGs were significantly detected (*p*<1×10^-8^) across all samples. No SNPs at the CpG, probe or single bases extension were annotated to the 15 *ACE2* CpGs. DNAm methylation was estimated on the β-value scale, from 0-1, reflecting the proportions of DNA molecules that are methylated at the target CpG.

#### Epigenetic Aging Biomarkers

We calculated two epigenetic aging biomarkers utilizing the Horvath’s online calculator (http://dnamage.genetics.ucla.edu/): DNA methylation telomere length (DNAmTL)^38^ and the “skin and blood epigenetic clock.”^39^ We evaluated their performance with scatterplots and Pearson’s correlation coefficient (*r*). We modeled the acceleration of these measures resulting from regressing DNAmTL and the skin and blood epigenetic clock on chronological age. A positive value of DNAmTL adjusted for age (DNAmTLadjAge) would indicate that DNAmTL is longer than expected based on chronological age (thus marker of “younger” cells), while positive residuals from the skin and blood clock would indicate epigenetic age acceleration (thus marker of “older” cells). We modeled the DNAmTLadjAge and epigenetic age acceleration for the skin and blood clock to estimate associations independent of chronological age.

#### Statistical Analyses

We described our study sample using means, standard deviations (SDs), counts and proportions. We extracted the *ACE2* gene genomic annotation from Bioconductor using the *IlluminaHumanMethylationEPICanno.ilm10b4.hg19* annotation package and calculated means and SDs of DNAm at the selected CpGs in the overall cohort and stratified by sex. We estimated the overall Spearman correlation structure among CpGs and used boxplots to visualize differences in DNAm by sex. We tested differences in DNAm by sex, age, race/ethnicity, DNAmTLadjAge, and the residuals of the skin and blood clock using robust linear regression models on beta values (0-1) to account for heteroskedasticity or influential points. We estimated differences by sex adjusting for chronological age and estimated differences by age stratified by sex. We estimated differences by race/ethnicity (each other category relative to white, the largest group) adjusted for age and stratified by sex. Differences in epigenetic aging biomarkers were adjusted for race/ethnicity and stratified by sex. We used localized Manhattan plots with the correlation matrix across CpGs to display differences by race/ethnicity using coMET.^40^ We report findings as %-DNA methylation differences and adjusted for multiple comparisons for each model independently using a False Discovery Rate of 5% (FDR<0.05). All statistical tests were two sided and analyses were carried out using R, version 4.0.2 (www.r-project.org/).

## Data Availability

Raw data to generate figures and tables are available from the corresponding author with the appropriate permission from the Project Viva study team and investigators upon reasonable request and institutional review board approval.

## Acknowledgments and Funding

This work was supported by the US National Institutes of Health grants R01ES031259, R01HD034568 and UH3OD023286.

## Contributions

AC performed the statistical analyses and wrote the main manuscript. DRG and EO conceived the initial study and provided critical feedback on the manuscript. SLRS, JES, DLD, AAB and MFH provided critical input on analyses and interpretation of the data. All authors reviewed the manuscript.

## Data availability

Datasets generated and analyzed during the current study are not publicly available because we did not obtain consent for such public release of epigenetic data from participants. Raw data to generate figures and tables are available from the corresponding author with the appropriate permission from the Project Viva study team and investigators upon reasonable request and institutional review board approval.

